# Highly valued despite burdens: qualitative implementation research on rapid tests for hospital-based SARS-CoV-2 screening

**DOI:** 10.1101/2021.08.27.21262660

**Authors:** Jonas Wachinger, Shannon A. McMahon, Julia Lohmann, Manuela De Allegri, Claudia M. Denkinger

## Abstract

Antigen-based rapid diagnostic tests (RDTs) for SARS-CoV-2 have good reliability and have been repeatedly implemented as part of pandemic response policies, especially for screening in high-risk settings (e.g., hospitals and care homes) where fast recognition of an infection is essential, but evidence from actual implementation efforts is lacking.

We conducted a prospective qualitative study at a large tertiary care hospital in Germany where RDTs are used to screen incoming patients. We relied on semi-structured observations of the screening situation, as well as on 30 in-depth interviews with hospital staff (members of the regulatory body, department heads, staff working on the wards, staff training providers on how to perform RDTs, and providers performing RDTs as part of the screening) and patients being screened with RDTs.

Despite some initial reservations, RDTs were rapidly accepted and adopted as the best available tool for accessible and reliable screening. Decentralized implementation efforts resulted in different procedures being operationalized across departments. Procedures were continuously refined based on initial experiences (e.g., infrastructural or scheduling constraints), pandemic dynamics (growing infection rates), and changing regulations (e.g., screening of all external personnel). To reduce interdepartmental tension, stakeholders recommended high-level, consistently communicated and enforced regulations.

Despite challenges, RDT-based screening for all incoming patients was observed to be feasible and acceptable among implementers and patients, and merits continued consideration in the context of rising infections and stagnating vaccination rates.

## INTRODUCTION

With vaccination rates stagnating and the emergence of novel virus variants potentially escaping vaccine-induced immunity, comprehensive testing and rigorous contact tracing remain a cornerstone of controlling the ongoing COVID-19 pandemic. (1–4) The risks of undetected infections resulting in severe morbidity and mortality are especially pronounced in settings where high-risk populations are clustered (e.g., retirement communities and health care facilities), underscoring a need for systematic screening of incoming persons to such settings.(5)

The gold standard approach to detecting SARS-CoV-2 infections, Reverse Transcription Polymerase Chain Reaction (PCR), is often not feasible to implement, especially in settings where rapid turnaround time is critical but also in other situations where testing capacity is constrained.(2) Antigen-based rapid diagnostic tests (RDTs) have thus been proposed as a timely, accessible, and pragmatic complement for screening of asymptomatic persons.(1) In December 2020, the WHO issued an implementation guide highlighting that – despite achieving inferior sensitivity and specificity levels compared to PCR – RDTs have the potential to rapidly detect active SARS-CoV-2 infections directly at the point of care while being easier to perform and less infrastructure-dependent and expensive.(6)

In high-income settings, with the exception of pediatric screening (e.g., for respiratory syncytial virus), RDTs have rarely been used; prior to the COVID-19 pandemic, universal screening independent of symptoms has not been done in high-income countries (HICs). Implementation insights on RDTs more generally stem primarily from low- and middle-income countries (LMICs) and draw on experiences using RDTs for individual diagnosis and not infection control.(7) Pre-admission SARS-CoV-2 RDT-based screening, particularly in health facilities, has repeatedly been used in HICs over the course of the pandemic. Although evidence suggests that such screening programs could have a promising diagnostic yield,(5, 8–10) research outlining how these programs are implemented and scaled up, and how those involved experience or adapt implementation is lacking. Given this paucity of information, the WHO called for further evidence from large-scale rollout of RDTs for SARS-CoV-2 to continuously update implementation guidance.(6)

With this study, we fill a gap in the literature by outlining RDT-based screening implementation processes and experiences at a major German university hospital. We aim to provide lessons learned to guide similar implementation efforts.

## METHODS

### Setting

Heidelberg University Hospital (UKHD in its German abbreviation) is among the largest tertiary care hospitals in Germany with over 13,000 employees and serving over 100,000 inpatients and 1.3 million outpatients per year.(11) By the end of data collection (March 3, 2021), a total of 19,270 SARS-CoV-2 cases and 415 deaths had been reported in the region served by the hospital.(12)

### Intervention Design

To reduce the risk of nosocomial transmission of SARS-CoV-2, UKHD introduced a hospital-wide, RDT-based screening for asymptomatic patients coming into the hospital for elective procedures (incl. day clinics) prior to admission starting from October 2020. The screening program addressed patients who were not able to provide documentation of a negative PCR result within 48h prior to admission. In addition, depending on current infection dynamics and changing regulations within and across departments, visitors, external contractors, or translators, were similarly screened. By the end of data collection, more than 50,000 RDTs have been performed in over 30,000 individuals.

Exact implementation procedures for the screening (e.g., screening location) were developed at the discretion of individual hospital departments and day clinics. Members of the hygiene department trained staff assigned to screening (predominantly nurses, in some departments also including medical students and other medical personnel), who then trained peers in a snowball system. Testing competency was assessed post-training. Screening was performed in designated areas in the respective departments using the STANDARD Q COVID-19 Ag Test (SD Biosensor, Inc. Gyeonggi-do, Korea), which is an independently validated, instrument-free lateral flow assay for SARS-CoV-2 detection that can be performed at point of care and is one of two RDTs listed by the WHO under the Emergency Use Listing.(13) In instances of a positive RDT result, the respective patient was isolated and retested using a validated SARS-CoV-2 PCR.

Confirmatory PCR testing was not recommended as a routine practice in case of a negative RDT result. Patients who developed symptoms associated with a SARS-CoV-2 infection or were in contact with confirmed cases were tested using PCR during their stay at the hospital. **Figure 1** summarizes the RDT-based screening approach as intended by the hospital’s task force.

**Figure 1:**
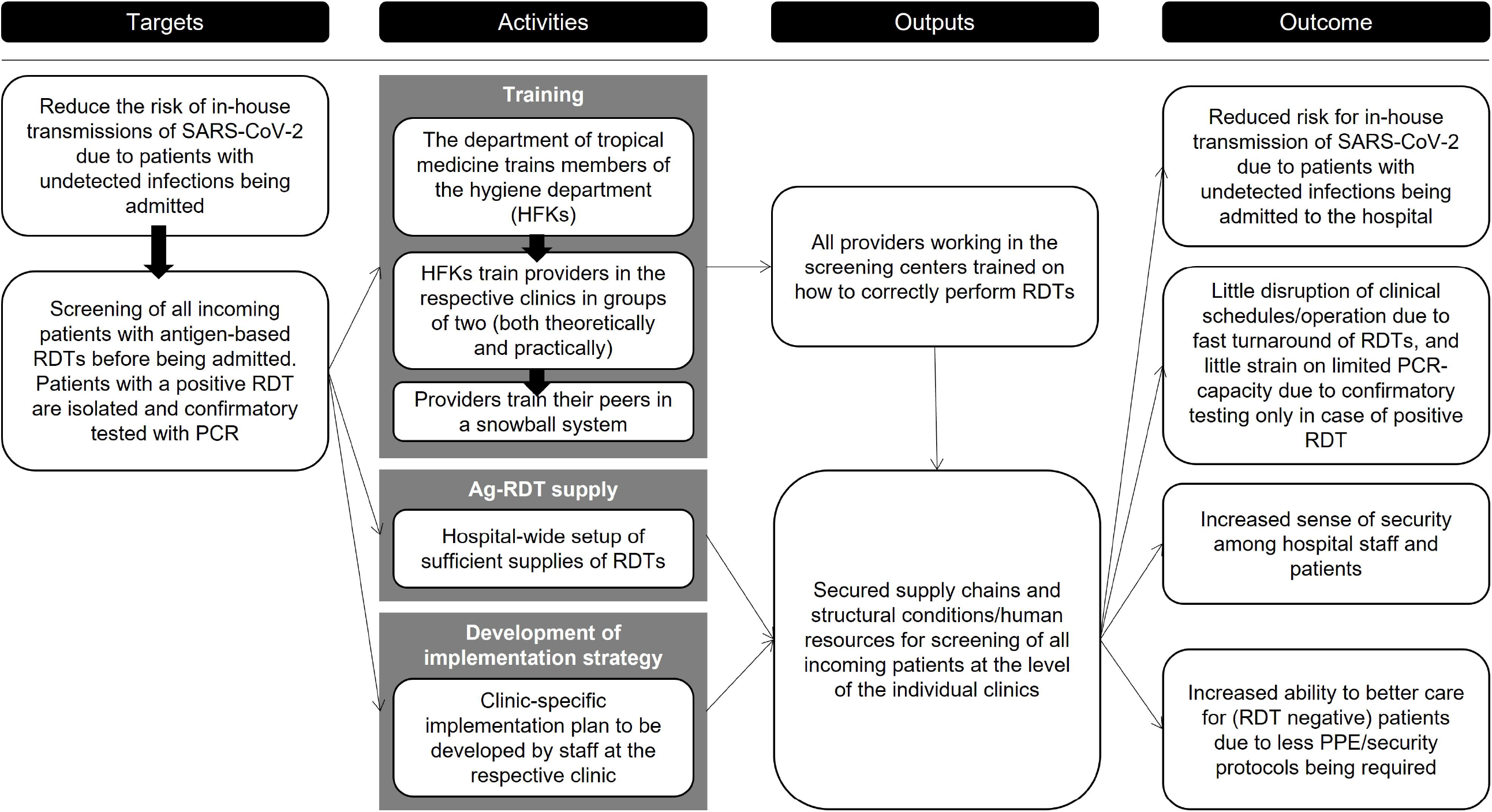
Theory of Implementation as intended by decisionmakers.

### Study procedures and sampling

To analyse implementation processes and experiences with RDT-based screening and drawing on the tenets of case study research(14) to develop an understanding of the issue from the perspective of participants, we conducted in-depth interviews with staff members at various hospital departments between November 2020 and March 2021. We purposefully selected a range of departments addressing different patient groups (children and adults) and clinical work (inpatient and outpatient). Respondent groups interviewed at the selected departments are outlined in **Figure 2**.

**Figure 2:**
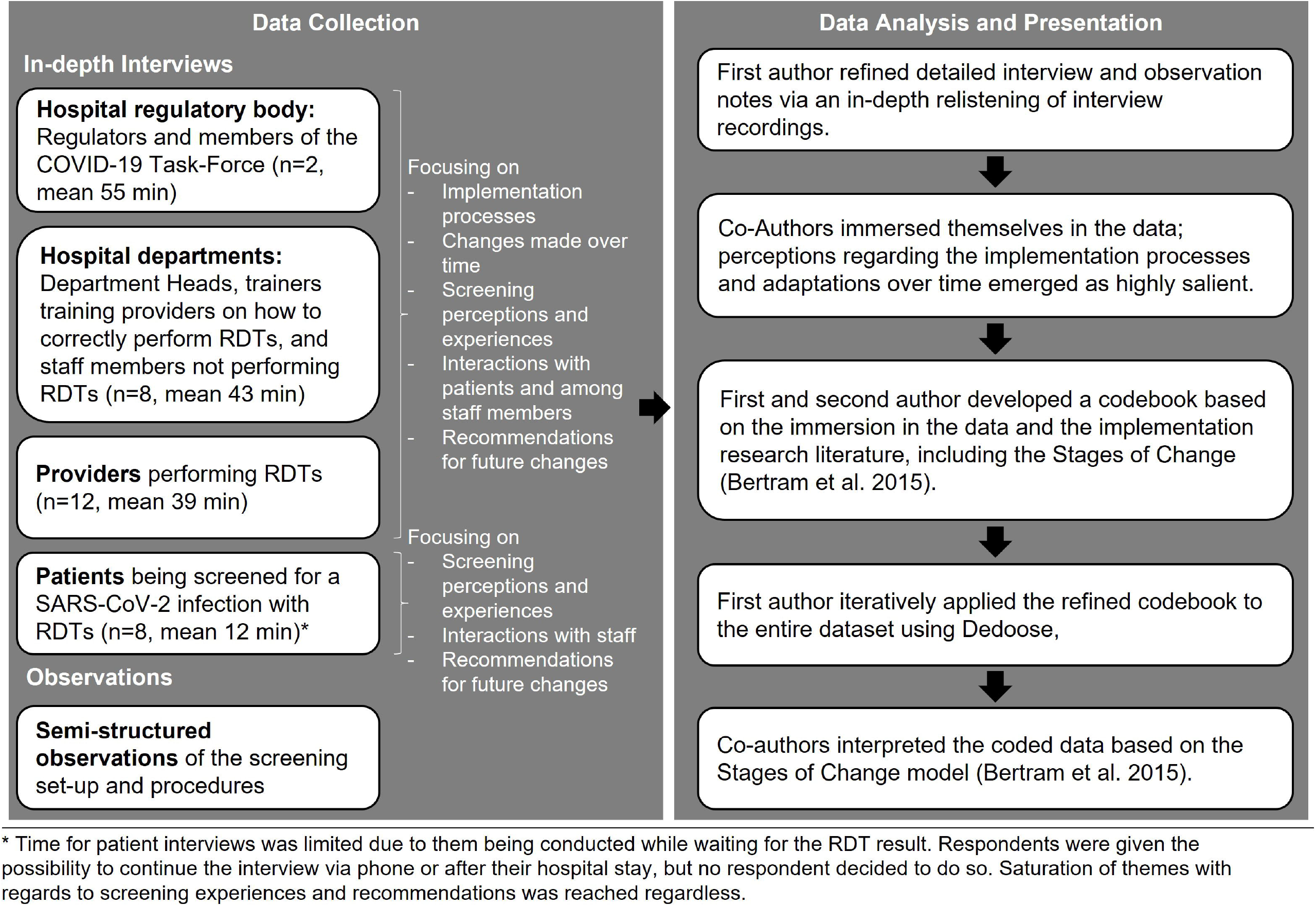
Summary of data collection and analysis processes.

Within each department and respondent group, respondents were purposefully selected and contacted through designated point persons at the respective department. For patient interviews, we used convenience sampling. We approached patients presenting at the RDT screening centre of the hospital’s medical clinic and conducted the interview while they awaited their test results. We drew on the COREQ-guidelines (see **supplemental file S1**) to report study procedures and findings. The ethical review board of the Medical Faculty, Heidelberg University, Germany (S-811/2020) approved this study. Prior to data collection initiation the study was registered in the German Clinical Trials Register (DRKS00023584).

### Data collection and analysis

In total, we conducted 30 interviews. Respondents provided written informed consent prior to interviews, and a majority of interviews were audio recorded (27), with three respondents objecting to audio recording. One department head objected to participation, quoting high pandemic-associated workload as the reason; 3 respondents did not acknowledge electronic invitations to participate. Two patients objected to participation, quoting unease with being interviewed in the context of waiting for their hospital appointment. The lead author, who has master’s level education in both Psychology and Medical Anthropology and extensive experience conducting qualitative interviews, conducted all data collection. Prior to each interview, the lead author introduced themselves and their role in the study, as well as the study procedures and objectives to establish a relationship and clarify study goals. The senior author developed the intervention with the task force.

Interviews were conducted in person in the clinic (for patients and a majority of staff members), with some interviews with staff members being conducted via phone or video call, according to respondent preference. The interviews were generally conducted with only the interviewer and respondent present. In the clinic context, due to respondent convenience and spatial constraints, sometimes non-participating individuals came into hearing distance; no participant preferred halting the interview until absolute privacy could be ensured.

The interviewer took detailed notes during interviews, and these notes were later supplemented with further information and quotes following an in-depth relistening of interviews. Interview questions, probes, and respondent categories were iteratively adapted based on ongoing dialogue within the research team.(15)

Once data saturation was reached, the lead co-authors immersed themselves in the data, discussing emerging topics and developing a codebook for analysis, focusing on implementation processes and experiences. The first author applied this codebook iteratively to the entire dataset. Processes and associated experiences, from the perspectives of the four main respondent groups outlined above, were mapped and interpreted by first and second author based on the stages of implementation as outlined by Bertram and colleagues’ model (Exploration, Installation, Initial Implementation, Full Implementation),(16) with the assistance of all co-authors (See Figure 2 for an outline of data collection and analysis processes). To protect respondent anonymity in the hospital setting, we attribute quotes based on respondent type only. One co-author was included as a respondent due to their role as a key stakeholder; no verbatim quote was included from this person.

## RESULTS

### Exploration, installation, and implementation of the RDT-based screening

Table 1 outlines our results following the four stages of implementation(16) and the four main respondent groups of this study as introduced above. We then outline implementation processes and experiences of the RDT-based screening, and how these changed over time, for each respondent group, highlighting heterogeneous experiences within and across respondent groups. Particularly salient themes are exemplified as key quotes in Table 2, as well as in the Case Studies 1 and 2.

**Table 1.**
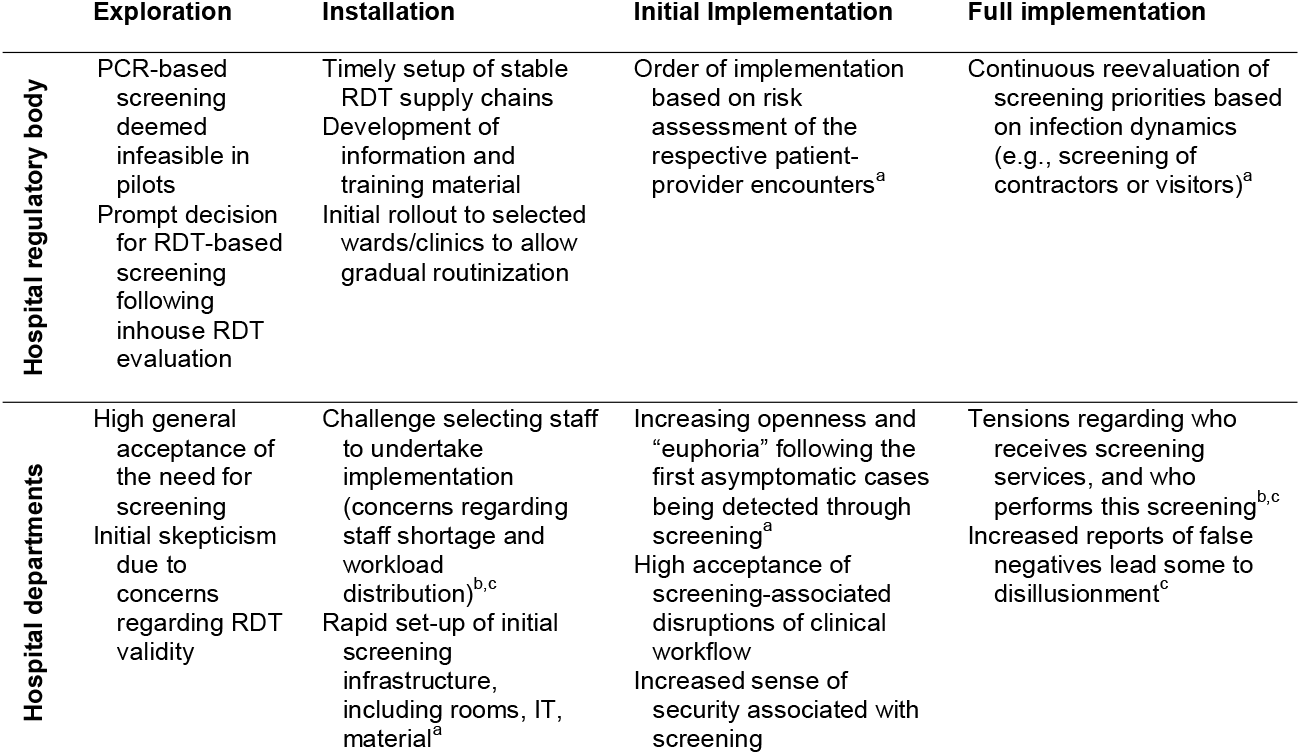
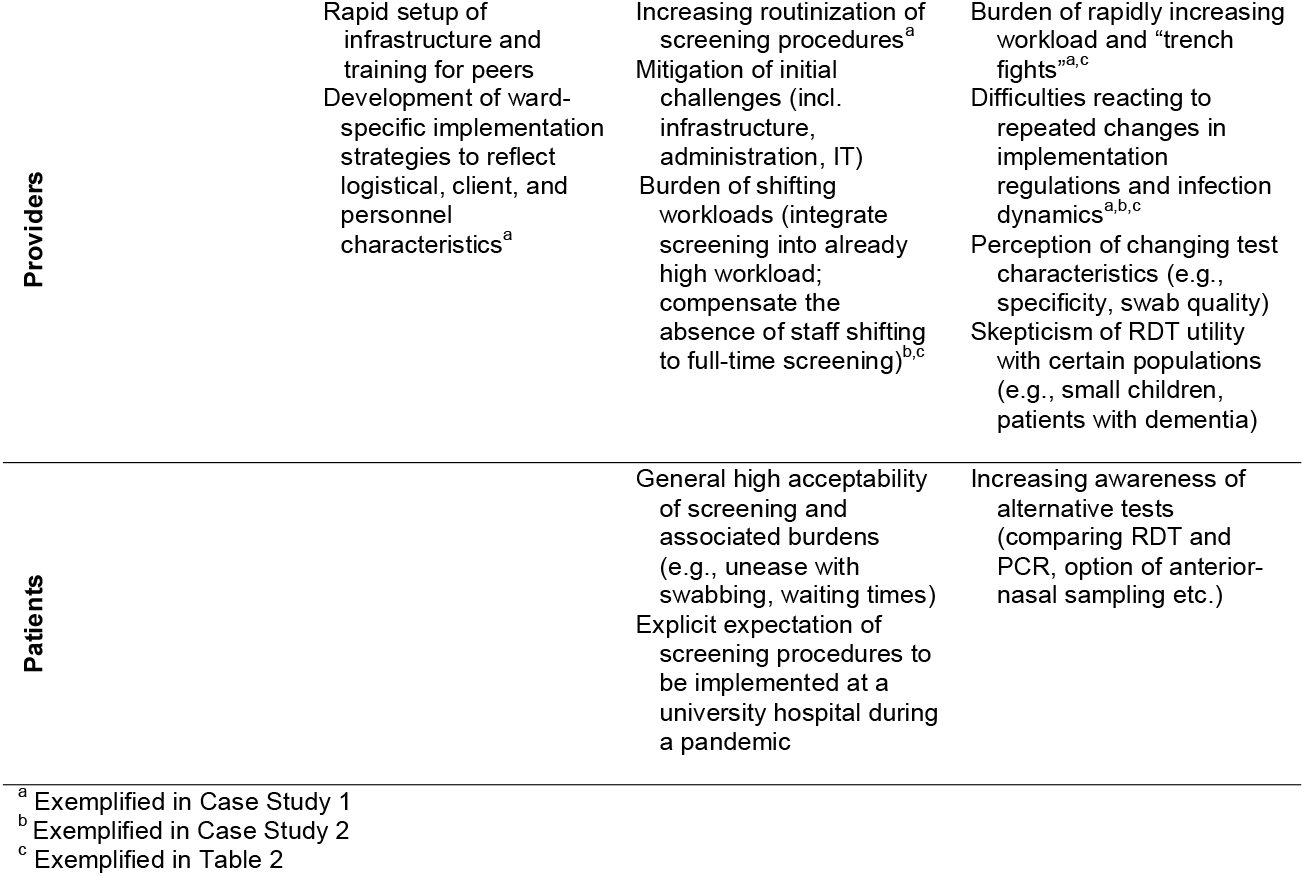
Exploration, Installation, and Implementation of the intervention.

**Table 2.**
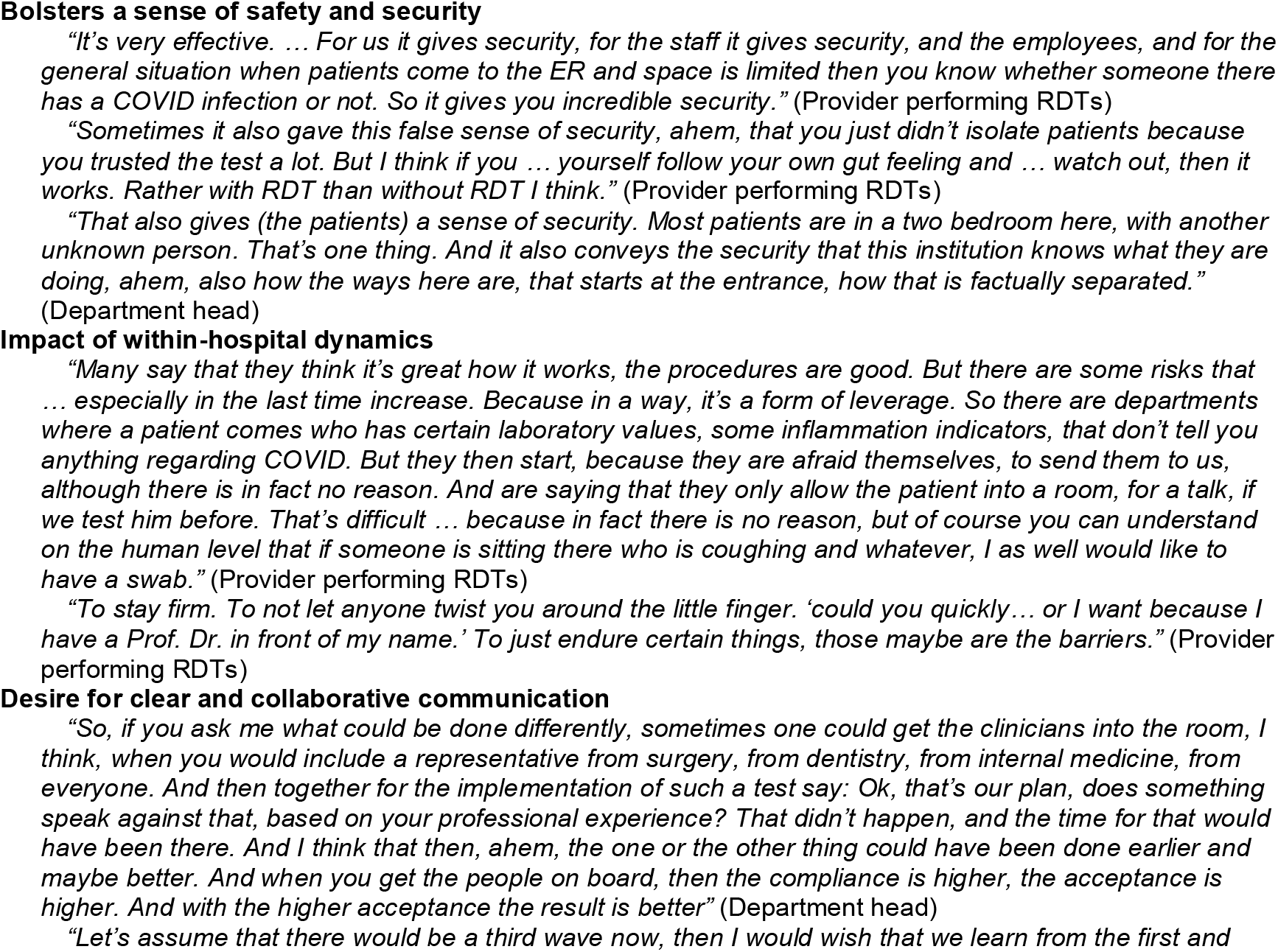
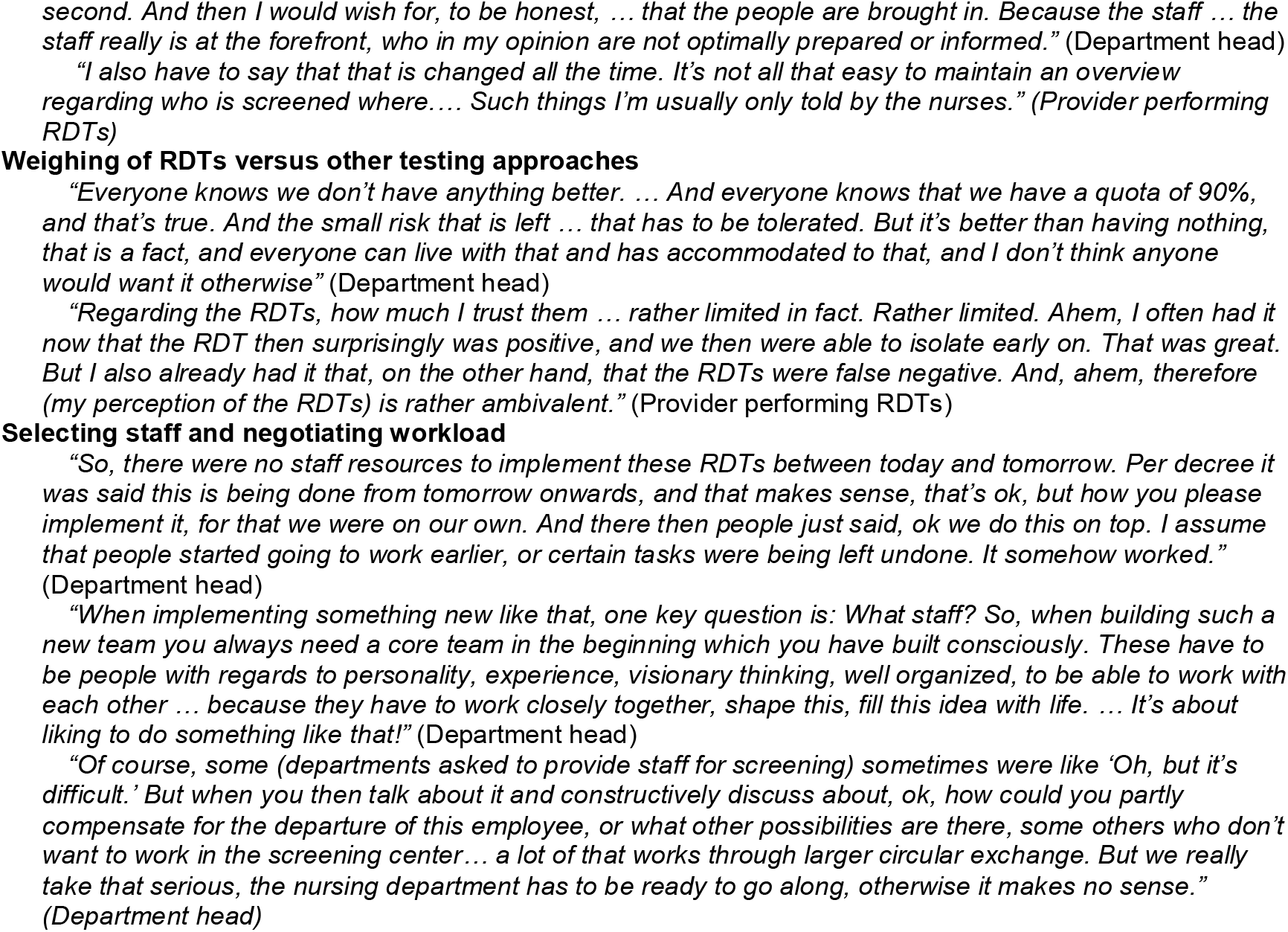
Acceptance of and experiences with RDT-based screening - illuminating quotes.

#### Members of the hospital regulatory body

The hospital regulatory body played a central role in the exploration and installation phases of the intervention. Respondents recounted how they initially had hoped for PCR-based screening to be feasible. However, due to laboratory constraints and a pilot of PCR screening in one ENT department which proved to be highly disruptive to clinical workflows, RDT-based screening was seen as the alternative “on the table” to contend with a second wave of infections. As large-scale validation studies of RDTs for SARS-CoV-2 were performed in the infectious disease department of the university hospital (a “fabulous situation for us”, Member of regulatory body), respondents recounted how they felt “encouraged” and had little objection to RDT-based screening once a test was identified that seemed to fit their main criteria: ease of use, rapid turnaround times (facilitating point-of-care administration and minimal staff training), consistent supply chains, and an acceptable sensitivity and specificity.

Once this test was identified in August 2020, the hospital regulatory body set up stable supply chains and developed overarching guidelines for screening implementation. Respondents said they did “not want to make any regulations for things we do not understand”, so exact implementation parameters were left to the discretion of individual departments who “are the ones on the ground and who know this”. At the end of September 2020, installation began “just in time” (Member of regulatory body) so that the screening could be adopted and refined prior to future infection waves. Regulators viewed their later role largely as providing refinements of the general regulations or guidance in light of new infection or patient dynamics.

#### Department heads and staff members

At the level of hospital departments, all respondents described a high acceptance of screening based on previous experiences with asymptomatic patients entering the hospital: “We knew what it feels like when cases are discovered only later… And that always resulted in this huge wave of contact tracing, contacts with staff, notifying the Gesundheitsamt [German regional health authority], sending patients or staff into quarantine and so on” (Department Head). One respondent described how she perceived such a rapid implementation only to be feasible because “I don’t think there was anyone who didn’t think this was a good thing” (Trainer). Some respondents reported that they themselves or colleagues initially were highly skeptical with regards to the reliability of RDTs and the potential disruption to clinical workflows induced by incorrect results but based on the recent in-house validation data most agreed that “that doesn’t look too bad – not 100%, but we can use that” (Department Head).

The selection of suitable location and personnel was among the core installation tasks for department heads. The selection of staff focused on an even distribution of burden across departments and identifying individuals who would be creative and nimble. Selected personnel were given a certain flexibility to develop procedures, “because they are the ones who actually work there and bring in their expectations and ideas regarding the organization (of the implementation). So, they shaped this, and they also reshaped it after some time to improve it even further. But that really was something we allowed the team to decide” (Department head). Certain wards whose patient groups did not qualify for screening by the centralized departmental screening centers (e.g., wards serving outpatients not requiring close contact treatment) decided to introduce screening independently, because “although integrating it into a normal schedule is hardly possible”, the screening was perceived as a “lesser evil” compared to the risk of nosocomial infections (Department head). Some participants said reports of false-negative RDT results caused concern or disillusionment. However, after roughly two months, the screening and its necessity was seen as generally highly accepted across wards, to the point that “if you would stop it now, that would lead to a lot of unrest” (Department head).

Communication between wards emerged as a key factor across interviews. Initially the screening resulted in some disruption of workflows on admitting wards. Over time, processes were adapted to minimize such disruptions, both on the side of the admitting wards (e.g., by informing patients to come earlier) and the respective screening center or personnel (e.g., by starting screening hours earlier). Despite continuing refinement of screening procedures and increasing acceptance, tensions between departments still emerged as some departments requested that ineligible patient groups be screened (e.g., ambulatory patients outside of day clinics).

#### Providers performing RDTs with patients

Most providers appreciated the flexibility they had regarding implementation processes and recounted how their daily experiences facilitated continuous implementation refinement: “Everything that has to do with humans you can plan on paper somehow and it then doesn’t work when the people come. And that’s how it works” (Provider performing RDTs). Several respondents recounted challenges of integrating the screening into an already high workload, both for themselves and for colleagues on the wards who had to compensate for screeners’ absence, and of contending with initial administrative and infrastructural challenges. Respondents also described how screening processes became more and more routinized, making it possible to accommodate rapidly increasing numbers of patients and personnel coming for screening: “You get used to everything, that’s the way it is here in the hospital” (Provider performing RDTs). The initially envisioned possibility for patients to provide proof of a recent negative PCR result as an alternative to the RDT-based screening was not reported as enacted by any department because confirming test documentation proved infeasible or outdated. Providers reported the test itself to generally be easy to perform, although testing small children (especially those aged 2-4) and patients with mental illness proved highly challenging.

According to several respondents, the most powerful and decisive factor in swaying teams to favor screening with RDTs was the discovery of an asymptomatic positive case: “So I remember, when the first positive patient was found, everything up there was standing still. Because no one had really expected that, because of course you think you have symptoms, you cough, you have fever. But these really are people who are, in quotation marks, completely healthy walking in here and are discovered by chance by this test” (Provider performing RDTs). As a result, several respondents described how the screening gave them an “incredible security” (Provider performing RDTs).

An additional burden to providers performing screening were “trench fights” associated with staff from various departments repeatedly trying to procure screening for ineligible patients, which providers described as “madness”, and something you had to “just endure” (Providers performing RDTs). Respondents shared how they appreciated their own department head’s support but would prefer enforcement of existing regulations on the level of regulatory bodies to minimize strain associated with repeated interdepartmental negotiations. Providers recounted how they often were surprised by the sudden, urgent need to implement novel screening strategies and to contend with constantly changing regulations. While they acknowledged that infection dynamics were fluid and regulations had to be adapted accordingly, several respondents described confusion regarding strategies that seemed to “not make sense”, be “stressful”, “not fair”, or “just impossible” (Providers performing RDTs) such as the screening of visitors allowed to enter the building, while at the same time denial of regular staff screening.

#### Patients screened with RDTs before being admitted to the hospital

Patients expressed high acceptance of screening, viewing it as “expectable” (even when they had not been forewarned), “essential”, “smooth”, and “for the benefit of everyone” (Patients screened with RDTs). One mentioned that their trust in the hospital would be diminished if “they weren’t doing something like this here … I can hardly imagine having trust in a hospital that doesn’t consider screening patients” (Patient screened with RDT). Some respondents compared the RDTs used in this setting with what they had experienced or heard about elsewhere, including test reliability (as compared to PCR), and sampling procedures, but generally acknowledged that the decisionmakers at the hospital would “know more about this”. Patients’ overall high acceptance was confirmed by providers, who described critical or rejecting voices being the minority and largely linked to misgivings about the swab itself or to exceptionally timely circumstances (such as rushing to see a dying relative). Patients voiced criticism regarding imprecise communication, regarding the value and duration of screening.

### Key implementation experiences across respondent groups

Experiences that were shared and reiterated across all respondent groups include the perception of increased safety associated with the screening; within-hospital (or cross-departmental) dynamics that could challenge implementation success; a desire for more timely information and collaborative decision making as programs are introduced or regulations change; the relevance of selecting engaged staff and fostering positive team dynamics; and regular weighing of RDTs versus other testing approaches (see **Table 2** for key quotes across each theme).

### Variation in adaptation between wards and over time

Implementation approaches and processes were continuously refined and varied across departments, based on departmental demands and workflows (e.g., patient characteristics, available space and personnel), changes in the guidelines of the regulatory body (e.g., based on changing infection dynamics or due to new insights etc.) or internal dynamics (e.g., demands of staff or patients, financial and infrastructural considerations etc.). To highlight the variability of intervention adoption and adaptation, we present two case studies: Case 1 is a centralized screening center responsible for screening patients admitted to several departments; Case 2 is a day clinic that repeatedly changed screening procedures for its outpatients over time based on patient and provider demands, changing infection dynamics, and the experience of nosocomial SARS-CoV-2 infections introduced by an asymptomatic outpatient.

#### Case Study 1. “It was a long process … but it worked well because everyone brought their ideas to the table” – A centralized screening center in a large clinic

Several departments opted to centralize RDT-based screening in one central area within the hospital’s main atrium. Providers working in this screening center described how the implementation process was challenging to organize and had to happen rapidly, but also said that a lack of rigid implementation regulations allowed for creativity. The existence of screening left several respondents with a sense of “supporting somewhere, where help is needed right now” (Provider performing RDTs. At the outset, only patients admitted to high-risk wards were systematically screened; this restricted roll-out was meant to limit screening burden and allow procedures to routinize prior to scale up. Within three months, screening expanded, and providers said that their routines had become “effective” and “like an assembly line”. While increases in the number of patients to be screened was “exhausting,” mutual reassurance bolstered a sense of teamwork: “We kept internal statistics, like ‘Oh, today we had 168, that’s the new house record!’ Also to show ourselves how much we had accomplished” (Providers performing RDTs).

Respondents described how departments often undervalued the effort required to screen. Beyond the swabbing itself -- the “easiest part” -- providers recounted how running the screening center also required complex documentation procedures and diplomatic skill when managing the queue during “rush hour” (Providers performing RDTs), factors left unacknowledged by some members of the hospital staff. One respondent described how she felt that screeners had become de facto first responders for individuals entering the building. Respondents described interactions with patients as predominantly positive albeit more perfunctory than either side would like: “In the beginning because of the just 20 patients per day we had a lot of time to talk with them, but by now you barely talk with the patients. You swab the patient and let them wait, and then the result is declared, and that’s it” (Provider performing RDTs). Respondents struggled to implement rapidly changing regulations regarding who gets screened. Declining screenings for overly eager wards or patient groups who did not qualify for screening was particularly “draining”. As one respondent said, “No one wants to say, ‘It’s too much, or it’s getting to be too much.’ But there are situations here and there where you feel you’re being used… (Department-based doctors) think, ‘Oh, it’s not too much. … I’ll just send this entire group to the screening center.’ … You really have to push back or you’ll get mowed over.” (Provider performing RDTs).

#### Case Study 2. “It changes all the time” – Continuous refinement of screening of outpatients who come for regular day clinic procedures

Even before the introduction of RDT-based screening, one department pre-emptively started weekly PCR testing of outpatients (all high-risk outpatients visiting multiple times a week). Despite this measure, one asymptomatic patient who had received a negative PCR result days prior, sparked infections among staff and patients. When RDTs became available hospital-wide, the department adopted the approach, but noted that RDTs required much more personnel time compared to PCR testing, which could be outsourced to a lab. “The additional work was huge. And until everyone was able to do that, let’s say handling-wise, that did take a little while. By now everything works very very well, but the initial time was challenging” (Department head).

Over the course of implementation, adaptations were frequently made to react to changing infection dynamics, requests from staff and patients, and infrastructural and financial circumstances: “In the beginning it was just when something was suspected. … And then it was once per week, then twice per week, then it was again reduced to once per week, and then we switched to this once PCR and once RDT” (Provider performing RDTs). Adaptations also included testing of patients before they entered the treatment room, or short-term increases in test frequency. These decisions were based on patients’ risk profiles. The latest screening approach of one PCR (beginning of the week) and one RDT (end of the week) was described as highly accepted by providers, both among their peers and patients.

## DISCUSSION

In this study, we outline implementation processes and experiences with RDT-based universal screening for SARS-CoV-2 in a tertiary care hospital setting in Germany. The screening was highly accepted across respondent groups. RDTs were described as imperfect regarding their reliability, but as the best available tool to facilitate entrance screening for all admitted patients. Implementation processes highlight how decentralized screening allows for setting up efficient workflows, but clear and, where possible, consistent instructions and regulations on screened patient groups and comprehensive communication of these regulations would reduce burden associated with interdepartmental negotiations

To the best of our knowledge, this is the first study to provide in-depth qualitative insights into the implementation of RDT-based screening for SARS-CoV-2, hence findings could inform implementation policies and processes in similar settings. However, the setting of a large university hospital and the fast-changing nature of the ongoing pandemic merits caution when making large-scale comparisons between groups and over time. Additionally, some findings and recommendations regarding implementation policies and procedures might not be applicable to other settings, including smaller or lower-resourced hospitals where staffing or test procurement challenges are more acute.

Both modeling(8, 9) and prospective studies(5, 17) have highlighted the potential of frequent and repeated screening for SARS-CoV-2. A number of authors and organizations, including the WHO, have repeatedly called for a rapid scale-up of RDT-based SARS-CoV-2 screening in various settings (e.g. hospitals, nursing homes, restaurants, or airports).(1, 2, 6, 18) Our study complements this ongoing discussion by highlighting that implementing RDT-based screening in a hospital setting is highly accepted albeit challenging.

Experience with RDT-based screening is limited to a few exceptions where point-of-care screening for influenza was introduced,(19) which commonly focus on assessing the diagnostic yield of a given screening. A scoping review assessing rapid, point-of-care testing for HIV in Canada found high acceptance and satisfaction across diverse population groups and screening locations (including hospitals).(20) Similarly, a systematic review of implementation barriers to point-of-care testing for HIV highlighted challenges such as a disruption of clinical workflows, which is mirrored in our study.(7) However, the employment of RDT-based screening for facility-wide infection control in a high-income setting in the case at hand presents a different use-case that limits comparability.

Our respondents appreciated clear regulations and logistical support, but emphasized how flexibility and more lead time were needed to develop context-specific working routines. This slightly contradictory finding (respondents want comprehensive updates but also fewer changes) highlights a challenge for policymakers to develop clear guidelines while responding to emerging, context-driven changes. Providing comprehensive blueprints for RDT-based screening that allow for adaptation to circumstances on the ground would be recommended across settings. Our study also highlights how conflicts arose due to attempts by some to procure screening for groups deemed ineligible by the regulator. Collaborative decision making, the enforcement of regulations at all levels, and clear attribution of roles and responsibilities would likely reduce interdepartmental negotiation.

## CONCLUSIONS

In summary, this study provides evidence for policymakers on how rapid universal screening at the point of care is feasible and highly accepted but requires clear guidance that can be adapted to local settings, as well as good communication on all levels. Considering the beginning fourth wave of infections in many countries and the emergence of vaccination-escaping SARS-CoV-2 variants, screening in high-risk settings will remain necessary. Further research is needed to determine the transferability of findings to other institutionalized settings such as nursing homes or schools, but also other settings marked by a set environment with stable groups of people and limited influx from outside, for example large companies. Additionally, both researchers and policymakers should consider how task-shifting to non-medical providers performing the tests could be optimized to minimize errors and maximize capacity. In general, point-of-care screening for infection control also has potential outside the pandemic context. Based on the results from our study, we encourage policymakers, researchers, and medical providers to consider exploring the potential of point-of-care screening for other infectious diseases (e.g., RSV and influenza) upon entering the hospital to reduce the risk of secondary infections within the hospital setting.

## Supporting information

Supplemental file S1 - COREQ checklist

## Data Availability

Considering the high public interest in research on COVID-19, qualitative data of participants who have indicated their agreement to this as part of the informed consent procedure can be shared with other researchers. However, to preserve the anonymity of respondents and considering the personal nature of qualitative data, requests will be considered on a case-by-case basis. Please contact the corresponding author.

## ACKNOWLEDGMENTS

We thank all participants for their time and contribution.

## SUPPORTING INFORMATION

S1 COREQ checklist. COREQ checklist for reporting qualitative research.

